# Assessing HIV vulnerability among young women in South Africa: A comparison of three risk assessment tools

**DOI:** 10.64898/2026.10.28.26366706

**Authors:** S. Dubazana, C. Hendrickson, R. Greener, M. Shahmanesh, D. Govindasamy, E. Musenge, J Miot, L. C. Long

**Author notes:** Corresponding author, Siyabonga Dubazana.

## Abstract

**Background:** Young women in sub-Saharan Africa remain disproportionately affected by HIV. However, self-perceived HIV risk may not align with reported sexual behaviors or other dimensions of risk. Behavioral measures, such as the Balloon Analogue Risk Task (BART), may capture risk-taking propensity not reflected in conventional self-report measures. We compared self-perceived HIV risk, behavior-based HIV risk, and behavioral risk-taking propensity among young women in KwaZulu-Natal, South Africa.

**Methods:** We conducted a cross-sectional survey between July and November 2025 among self-reported HIV-negative women aged 18–30 years in KwaZulu-Natal, South Africa. We assessed self-perceived HIV risk using the Perceived Risk of HIV Scale (PRHS), behavior-based HIV risk using the Vaginal and Oral Interventions to Control the Epidemic (VOICE) Risk Score, and behavioral risk-taking propensity using the Balloon Analogue Risk Task (BART). Spearman’s rank correlation coefficients were used to examine correlations between the three measures.

**Results:** Among 250 participants, median age was 22 years (IQR 20-25), all identified as Black, and 53.6% were attending school or college. Overall, 42.0% were classified as high risk by VOICE, 6.0% by PRHS, and 33.1% of participants with complete BART data were in the highest tertile of risk-taking propensity. Mean age differed across VOICE categories (*p* = 0.036), but not PRHS (*p* = 0.111) or BART categories (*p* = 0.698). PRHS categories were associated with anal intercourse, oral intercourse, consistent condom use, ever having sold sex, and age-disparate partnerships (all *p* < 0.05). VOICE categories were not significantly associated with the assessed sexual behaviors, while BART categories were associated with vaginal intercourse in the past month (*p* = 0.042). Correlations between PRHS, VOICE, and BART were weak and non-significant (Spearman’s ρ = −0.050 to 0.038; all p > 0.05).

**Conclusions:** These findings highlight the importance of distinguishing between reported HIV risk behaviours, perceived HIV risk, and behavioural risk-taking propensity when assessing HIV vulnerability among young women. For HIV prevention programmes, reliance on a single measure may not adequately capture the different dimensions of vulnerability. Integrating behavioural assessments with discussions of perceived risk and individual circumstances may help inform more responsive, person-centred HIV prevention services, including PrEP counselling. Further research is needed to establish how these measures can best complement one another in prevention service delivery.

## Introduction

Globally, an estimated 40.8 million people were living with HIV in 2024, with 1.3 million new infections, over three times the 2025 The Joint United Nations Programme on HIV/AIDS (UNAIDS) target of 370,000 (1). Sub-Saharan Africa (SSA) remains the epicentre of the HIV epidemic, accounting for approximately 25.6 million people living with HIV and 640,000 new HIV infections annually, almost half the global total (1). Despite these daunting figures, the region has seen a 56% reduction in new infections since 2010, largely driven by the scale-up of large national treatment and prevention programmes as well as donor led programs like Determined, Resilient, Empowered, AIDS-free, Mentored, Safe (DREAMS) (2-5). More recently, the introduction and scale-up of daily oral pre-exposure prophylaxis (PrEP) provision has added an additional prevention option, although its population-level impact remains modest due to variable uptake and adherence (6, 7).

Despite these promising gains, adolescent girls and young women (AGYW) in SSA continue to bear a high burden of HIV. In 2023, an estimated 3,100 AGYW were newly infected each week within the region, more than three times that of similarly aged males (8, 9). Multiple intersecting biological, behavioral and structural factors heighten vulnerability of AGYW to HIV; these can include biological susceptibility, early sexual debut, gender-based violence, transactional sex, and age-disparate relationships (10-16). Community-level drivers such as stigma and entrenched social norms, together with structural determinants like poverty, migration, and limited access to healthcare further compound young women’s vulnerability to HIV infection (17, 18). While these biological and structural drivers create an objective landscape of vulnerability, women’s engagement with HIV prevention tools also depends on subjective perception and appraisal of HIV susceptibility; these two dimensions of HIV vulnerability are often not aligned.

Evidence consistently shows a disconnect between self-perceived and behavior-based HIV vulnerability in SSA. For example, South African studies indicate that many young women underestimate their HIV vulnerability, even when engaging in high-risk behaviors (19, 20). This pattern is also observed in other SSA settings. In Western Kenya, a survey of AGYW aged 15-25 found an HIV prevalence of 4.5%, yet only 14.5% of sexually active participants reported high risk perception (21). Similarly, in Malawi, among AGYW with more than eight HIV-related risk factors, 52% did not perceive themselves to be at high risk, and 21% reported no HIV-worry at all (22). Traditional self-oriented risk-perception measures are therefore limited in identifying individuals who would benefit from prevention interventions. This limitation is also reflected in the performance of HIV behavior-based risk assessment tools. A recent systematic review indicates that validated behavior-based tools, including the VOICE and other multi-domain risk scores, demonstrate only modest predictive performance among cisgender women, including both AGYW and older women (23). Although these tools tend to have high sensitivity at commonly used cut-offs, their low specificity limits their ability to accurately distinguish who will acquire HIV from those who will not (24).

Traditional approaches to assessing risk perception often rely on single-question measures, which may inadequately capture the multidimensional nature of risk appraisal (25). Similarly, tools assessing behavioral risk are limited in that they only capture historical and self-reported behaviors, making them vulnerable to recall and social desirability bias, while potentially failing to reflect current circumstances or future risk trajectories.

Behavioural economics tools such as the BART, a validated computerized measure of risk-taking propensity, offer an alternative approach by measuring risk-taking propensity through observed decision-making, rather than self-report. This tool measures ‘impulsivity’ or ‘reward-seeking behavior’ which are behavioral phenotypes often associated with inconsistent condom use or multiple sexual partners (26, 27). The BART may bypass social desirability bias where participants might under-report their self-perceived vulnerabilities or past behaviors to avoid judgement. The BART has been validated in domains such as substance use and sexual risk-taking (27, 28), yet its application to understanding HIV-related decision-making among young women remains limited.

Understanding how behaviorally measured risk-taking propensity aligns with self-perceived and behavior-based HIV risk is crucial for identifying young women who may be most vulnerable yet underestimate their risk. By examining correlations between BART-assessed risk propensity, self-perceived HIV risk, and behaviour-based risk, this study provides innovative insights into the interplay between cognitive, behavioural, and self-reported risk factors in a rural, high-HIV burden setting. In light of this, our study aimed to describe the relationships between BART-assessed risk propensity, self-perceived HIV risk, and behaviour-based HIV risk tools.

## Materials and Methods

### Study design, setting, and population

We conducted a quantitative, cross-sectional observational study between July and November 2025 in uMkhanyakude, KwaZulu Natal (KZN), South Africa, within the Africa Health Research Institute’s (AHRI) Health and Demographic Surveillance System (HDSS). The HDSS provides a longitudinal population-based research platform for monitoring demographic and health-related events among residents in the study area. uMkhanyakude is a predominantly rural area near the Mozambique border characterized by dispersed households, high HIV prevalence (36.3% among adults), and high youth unemployment (>85%) (29, 30). Recruitment was embedded within the ongoing *Thetha Nami, Ngithethe Nawe* (hereafter, *Thetha Nami*), a stepped-wedge cluster-randomized trial evaluating the effectiveness and implementation of a peer navigator-led, community-based intervention to reduce HIV transmission among young people aged 15–30 years in KZN (31-33).

### Sampling and recruitment

We purposively sampled potential participants from the *Thetha Nami* safe spaces (34). We conducted a power assessment using Fisher’s *z* transformation. With a sample size of 250, the study had 80% power to detect correlations of approximately *r*=0.18 or larger at a 5% significance level. This sample size was deemed adequate to examine associations between risk-taking propensity (BART), self-perceived HIV risk (PRHS), and HIV risk behaviours (VOICE risk score), while remaining feasible within the recruitment capacity of the *Thetha Nami*, safe-space sessions. Following the completion of a safe space session, potential participants were screened for eligibility. Participants were eligible if they were female sex-assigned at birth, aged 18-30 years, attending the safe spaces, self-reported as HIV negative or of unknown serostatus, and able to complete a self-administered survey in English or isiZulu on a handheld electronic tablet.

### Data collection

Following informed consent, participants completed a self-administered survey on electronic tablets (33). A self-administered, tablet-based approach was chosen to enhance privacy given the sensitive nature of some questions. The BART was implemented using oTree, an open-source platform for behavioural experiments (35). After completing all study procedures, participants received a ZAR100 (USD 6.02; 2025 average exchange rate) gift card as reimbursement for their time to participate.

### Self-reported HIV risk behaviours

We assessed behaviour-based HIV vulnerability using the VOICE Risk Score, which determines a score from 0-11 based on age, relationship status, financial or material support from partner, partners of primary partner, alcohol use, current sexually transmitted infection, and herpes simplex virus (HSV) type 2 status (36). The original score incorporates laboratory-based STI diagnoses and HSV-2 serostatus, however, these measures were not available in our study. The score was therefore adapted to reflect the variables available in our dataset, a limitation that may affect its comparability with the original VOICE Risk Score. This scale has been used in a South African context with AGYW and has been used in various SSA settings, including Malawi, Uganda, Zimbabwe, and Zambia with young African women (36, 37). The VOICE risk score is a composite epidemiologic index derived from multiple independent predictors of HIV acquisition. For this study, total scores were calculated for each participant and categorized into three risk levels: low risk (0-3 points), moderate risk (4-8 points), and high risk (≥9 points). As the score represents a formative construct rather than a reflective latent scale, internal consistency reliability (Cronbach’s alpha) was not considered an appropriate metric for evaluation.

### Perceived risk assessment

Self-perceived risk was assessed using the PRHS, an eight-item instrument capturing cognitive, affective, and salience dimensions of HIV risk perception (38). Though this scale has not been used in South Africa, it has been used in similar contexts such as Eswatini with adolescents and young adults and in Nigeria among school-going adolescents (39, 40). For descriptive and analytical purposes, PRHS scores were categorized as follows: low (8-22), moderate (23-33), and high (34-40) perceived risk. In our study, this 8-item scale demonstrated acceptable internal consistency (Cronbach’s α = 0.74).

### Risk-taking propensity

We measured risk-taking propensity using the Balloon Analog Risk Task (BART), a computerized behavioural tool that assesses willingness to take risks for potential rewards (41). In the BART,), participants inflate virtual balloons to earn points. Each pump increases the potential reward, but also exposes participants to the risk of the balloon bursting and losing the accumulated points for that balloon. Participants must therefore balance the potential gains from additional pumps against the risk of losing their accumulated rewards. (41). Participants must decide when to stop inflating and “bank” their points to avoid losing them. Each participant completed 20 rounds (one balloon per round), providing multiple opportunities to engage in risk-reward decisions. Points were virtual and were not exchangeable for real monetary rewards. Following the standard methodological convention for the BART the average adjusted number of pumps served as the risk-taking proxy, with higher adjusted-average pumps reflecting greater risk-taking propensity, whereas lower values reflect more risk-averse behaviour. Adjusted pumps were calculated by averaging the number of pumps in rounds where the balloon did not explode. This measure captures participants’ voluntary risk-taking behaviour, excluding rounds censored by the random explosion point, and thus provides a more accurate representation of risk propensity than an average across all rounds.

### Data analysis

We analysed these data using a cross-sectional quantitative design, using Stata Release 19 (42). We used descriptive statistics to summarize participants’ sociodemographic and behavioural characteristics. Chi-square tests were used to assess associations between categorical participant characteristics and categories of the three risk assessment measures. For correlation analyses, BART-adjusted average pumps, PRHS scores, and VOICE risk scores were standardized to z-scores. Spearman’s rank correlation coefficients (ρ) were then calculated to examine the relationships between behavioural risk-taking propensity, self-perceived HIV risk, and behaviour-based HIV risk. Spearman’s method ranks participants rather than using raw values, making it robust to differences in scale and skewed distributions. This non-parametric approach allowed for a standardized comparison of the three distinct dimensions of risk without requiring linear assumptions. For comparative analyses, the BART was categorized into tertiles (lowest, middle, and highest thirds) based on its distribution within the study sample. This approach addressed the absence of validated cut-off values for the BART, allowing participants to be grouped into approximately equal-sized categories representing increasing levels of risk. To assess the convergence of these measures, we conducted a multi-dimensional matrix analysis across three standardized measures: risk-taking propensity (BART), the VOICE Risk Score, and the PRHS Score (Figure 1). All statistical tests were two-sided, with significance defined as p<0.05. To facilitate direct comparison across the three HIV risk metrics, all raw scores were converted into a standardized Proportion Score (0-100%) (Figure 2). For VOICE and PRHS, we calculated this as the percentage of the maximum possible assessment tool score. For the BART, scores were standardized as a percentage relative to the maximum mean adjusted pumps observed within the sample. These proportion scores were used for descriptive analyses and visualization of score distributions across metrics.

**Figure 1.**
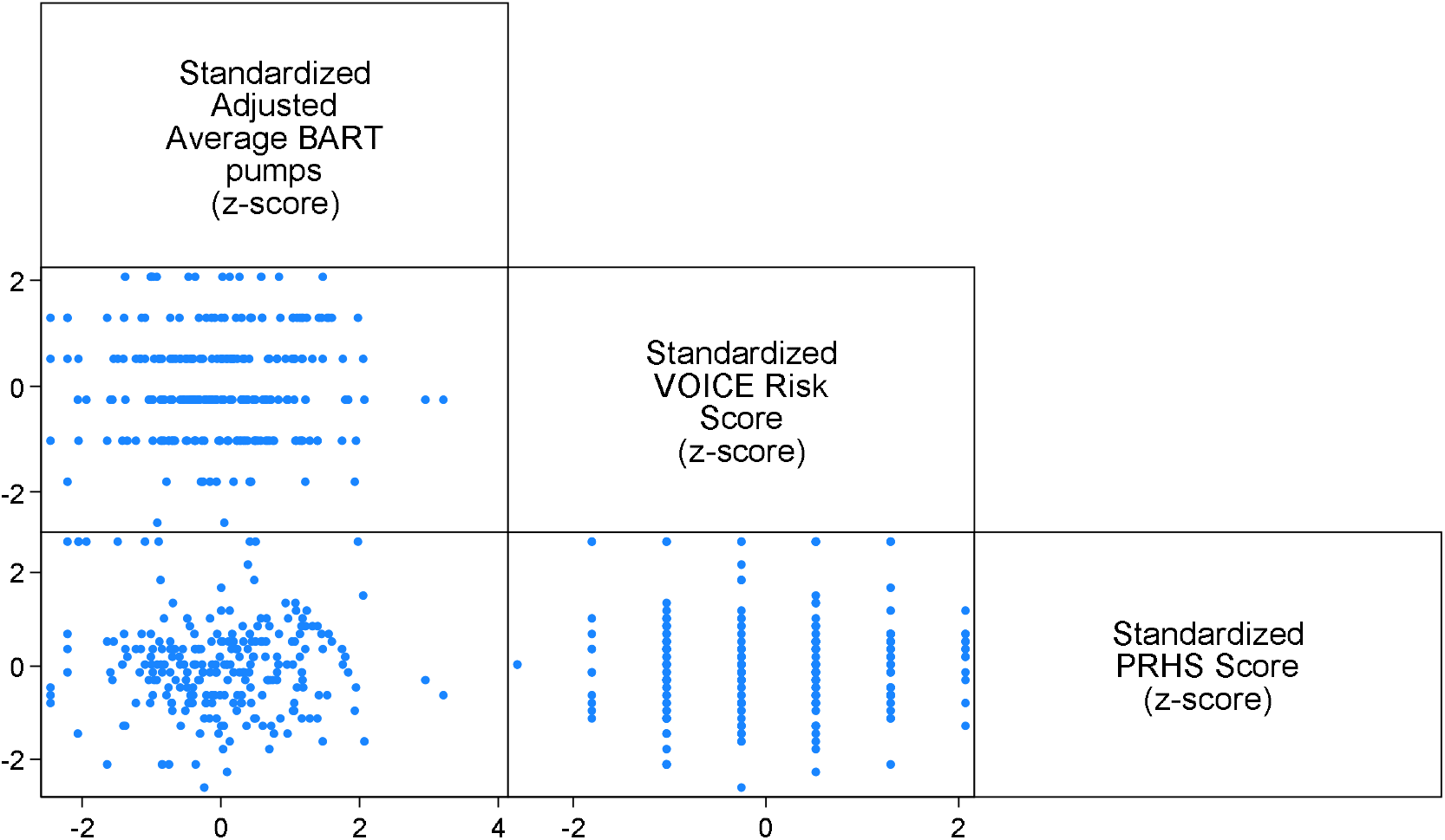
Correlation Analysis of Behavioral Risk Metrics.

**Figure 2.**
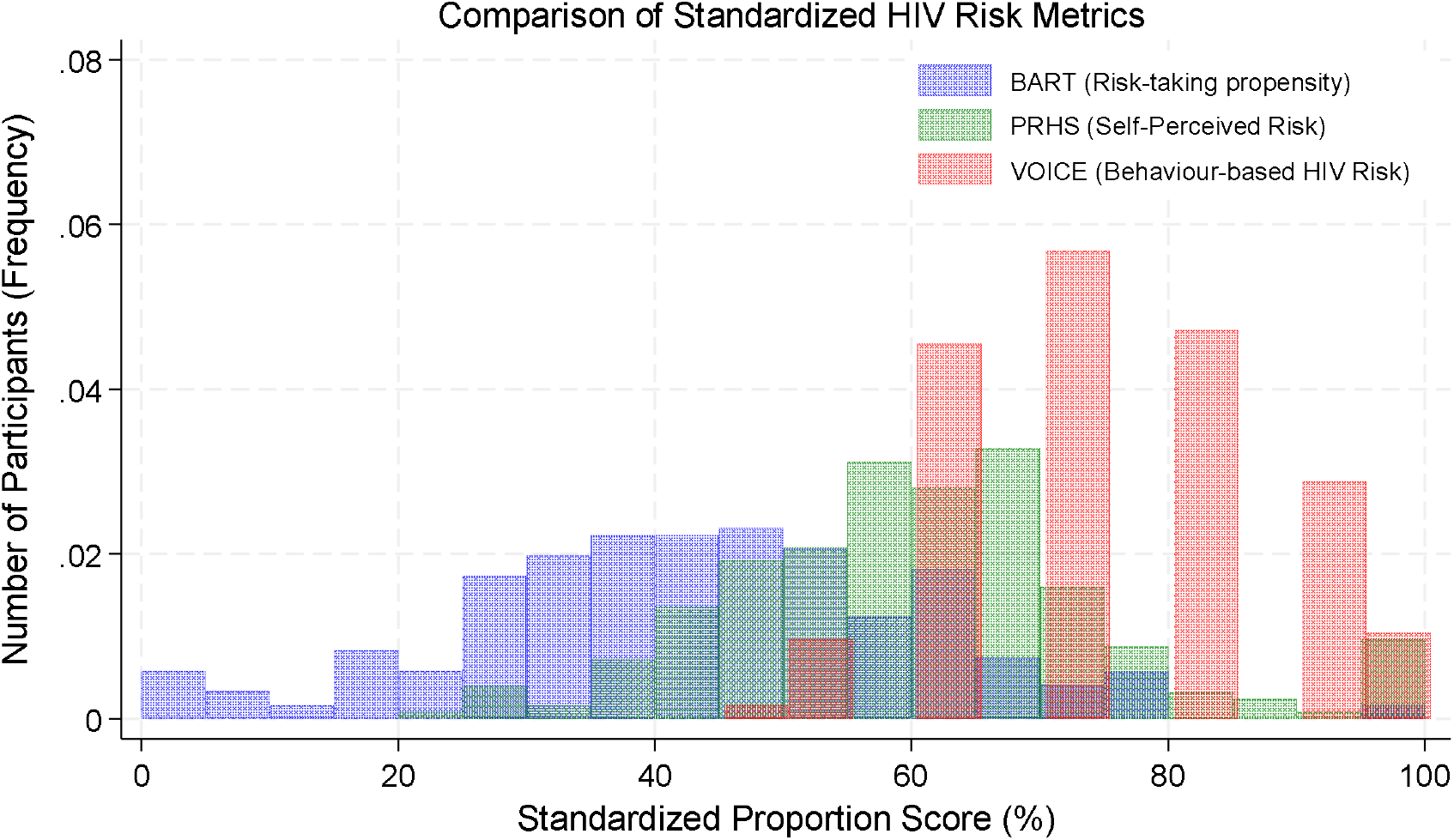
Distribution of Standardized Scores Across Assessment Metrics.

### Ethical considerations

Ethical approval was obtained from the University of the Witwatersrand Human Research Ethics Committee (M241008), the KZN provincial Department of Health and the uMkhanyakude District (NHRD ref: KZ_202408_029).

## Results

### Participant characteristics

Of the 256 women approached, 250 (97.7%) completed the survey. BART data were missing for eight participants, these cases were excluded from analyses involving BART, with the available data used for other analyses. Participants had a median age of 22 years (IQR: 20–25), all self-identified as Black, and just over half (53.6%) were currently attending school or college (Table 1). Socioeconomic constraints were common: 34.4% reported that their household sometimes or often went without food, while 59.6% reported difficulty or great difficulty accessing ZAR 100 for healthcare. Sexual and partnership characteristics varied across participants. Although most reported one sexual partner in the preceding six months (83.6%) and a primary partner at their last sexual encounter (93.6%), 56.4% reported having had sex with someone more than five years older. Condom use was inconsistent, with 43.6% reporting condom use at last sex and 42.0% reporting condom use during every sexual interaction in the preceding month. Nearly half (46.0%) did not know their partner’s HIV status, and 16.5% reported having ever sold sex.

**Table 1.**
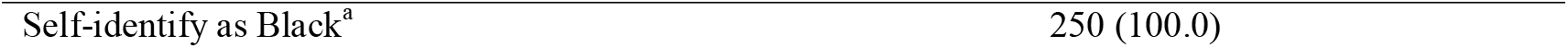
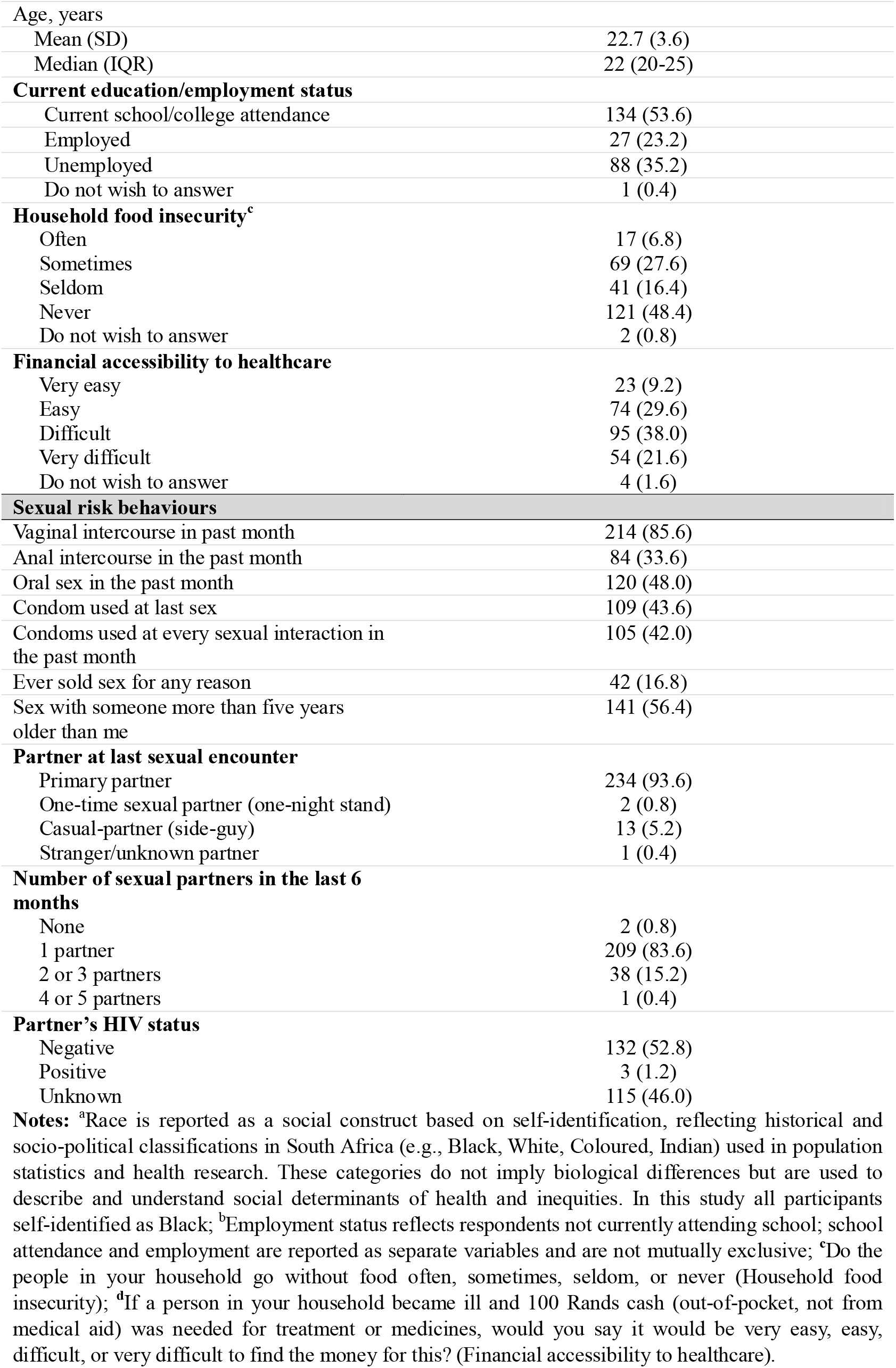
Sociodemographic, household, and sexual risk characteristics.

### Participant characteristics by VOICE risk score

Participants were most commonly classified in the moderate-risk VOICE category (54.4%), followed by the high-risk (42.0%) and low-risk (3.6%) categories (Table 2). Mean age differed significantly across categories (p=0.036), with participants in the high-risk category being younger than those in the moderate- and low-risk categories. Household food insecurity also differed across categories (p=0.029); however, participants classified as high risk more frequently reported never experiencing household food insecurity than those in the moderate- and low-risk categories. No significant differences were observed across VOICE categories in education/employment status, financial accessibility to healthcare, sexual risk behaviours, partner characteristics, or partner HIV status.

**Table 2.**
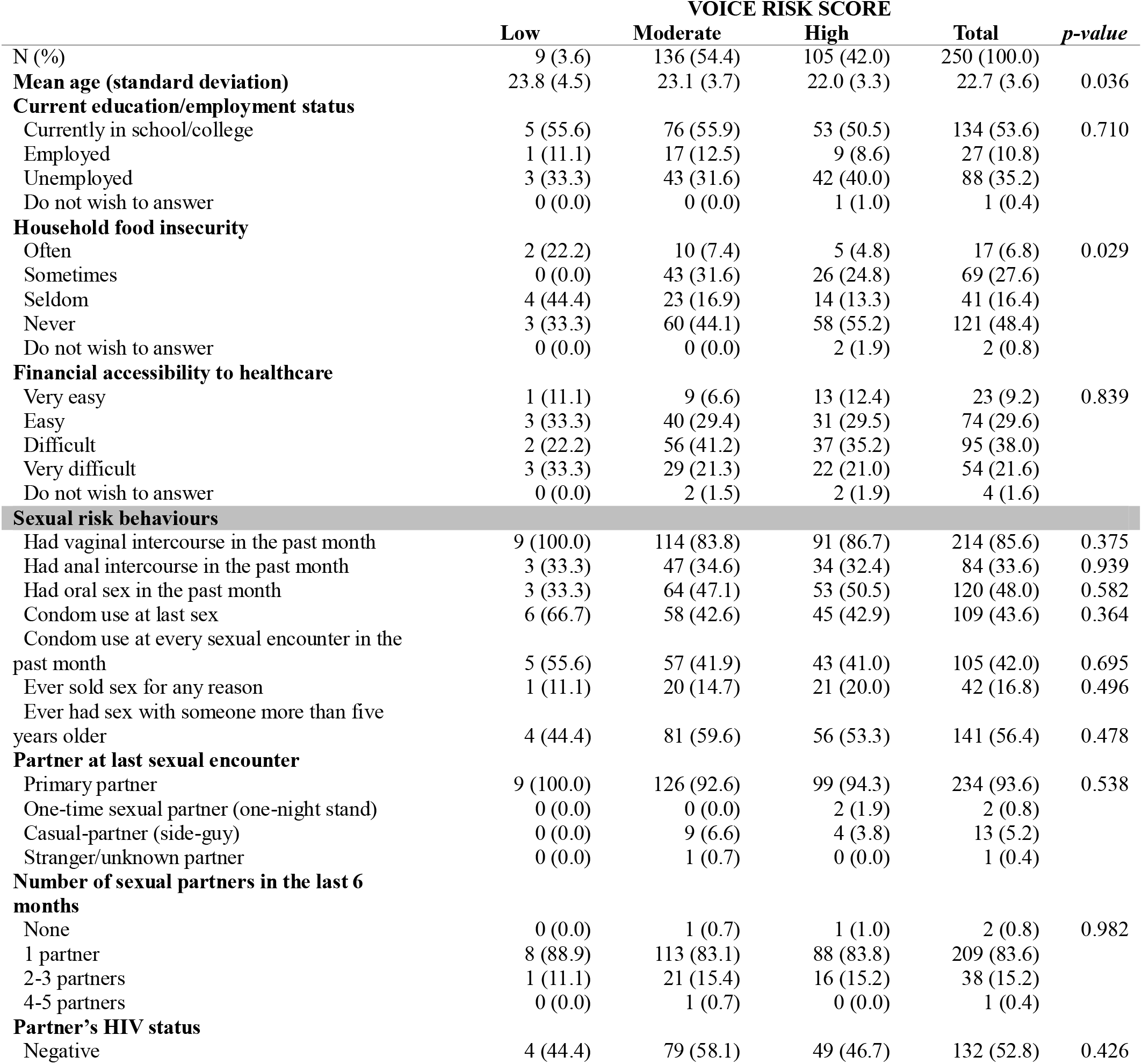
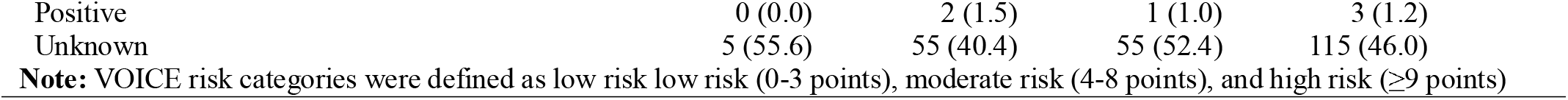
Sociodemographic, household, and sexual risk behaviour characteristics by VOICE Risk Score.

|  | VOICE RISK SCORE |  |  |  | *p-value* |
| --- | --- | --- | --- | --- | --- |
|  | Low | Moderate | High | Total |  |
| N (%) | 9 (3.6) | 136 (54.4) | 105 (42.0) | 250 (100.0) |  |
| **Mean age (standard deviation)** | 23.8 (4.5) | 23.1 (3.7) | 22.0 (3.3) | 22.7 (3.6) | 0.036 |
| **Current education/employment status** |  |  |  |  |  |
| Currently in school/college | 5 (55.6) | 76 (55.9) | 53 (50.5) | 134 (53.6) | 0.710 |
| Employed | 1 (11.1) | 17 (12.5) | 9 (8.6) | 27 (10.8) |  |
| Unemployed | 3 (33.3) | 43 (31.6) | 42 (40.0) | 88 (35.2) |  |
| Do not wish to answer | 0 (0.0) | 0 (0.0) | 1 (1.0) | 1 (0.4) |  |
| **Household food insecurity** |  |  |  |  |  |
| Often | 2 (22.2) | 10 (7.4) | 5 (4.8) | 17 (6.8) | 0.029 |
| Sometimes | 0 (0.0) | 43 (31.6) | 26 (24.8) | 69 (27.6) |  |
| Seldom | 4 (44.4) | 23 (16.9) | 14 (13.3) | 41 (16.4) |  |
| Never | 3 (33.3) | 60 (44.1) | 58 (55.2) | 121 (48.4) |  |
| Do not wish to answer | 0 (0.0) | 0 (0.0) | 2 (1.9) | 2 (0.8) |  |
| **Financial accessibility to healthcare** |  |  |  |  |  |
| Very easy | 1 (11.1) | 9 (6.6) | 13 (12.4) | 23 (9.2) | 0.839 |
| Easy | 3 (33.3) | 40 (29.4) | 31 (29.5) | 74 (29.6) |  |
| Difficult | 2 (22.2) | 56 (41.2) | 37 (35.2) | 95 (38.0) |  |
| Very difficult | 3 (33.3) | 29 (21.3) | 22 (21.0) | 54 (21.6) |  |
| Do not wish to answer | 0 (0.0) | 2 (1.5) | 2 (1.9) | 4 (1.6) |  |
| **Sexual risk behaviours** |  |  |  |  |  |
| Had vaginal intercourse in the past month | 9 (100.0) | 114 (83.8) | 91 (86.7) | 214 (85.6) | 0.375 |
| Had anal intercourse in the past month | 3 (33.3) | 47 (34.6) | 34 (32.4) | 84 (33.6) | 0.939 |
| Had oral sex in the past month | 3 (33.3) | 64 (47.1) | 53 (50.5) | 120 (48.0) | 0.582 |
| Condom use at last sex | 6 (66.7) | 58 (42.6) | 45 (42.9) | 109 (43.6) | 0.364 |
| Condom use at every sexual encounter in the past month | 5 (55.6) | 57 (41.9) | 43 (41.0) | 105 (42.0) | 0.695 |
| Ever sold sex for any reason | 1 (11.1) | 20 (14.7) | 21 (20.0) | 42 (16.8) | 0.496 |
| Ever had sex with someone more than five years older | 4 (44.4) | 81 (59.6) | 56 (53.3) | 141 (56.4) | 0.478 |
| **Partner at last sexual encounter** |  |  |  |  |  |
| Primary partner | 9 (100.0) | 126 (92.6) | 99 (94.3) | 234 (93.6) | 0.538 |
| One-time sexual partner (one-night stand) | 0 (0.0) | 0 (0.0) | 2 (1.9) | 2 (0.8) |  |
| Casual-partner (side-guy) | 0 (0.0) | 9 (6.6) | 4 (3.8) | 13 (5.2) |  |
| Stranger/unknown partner | 0 (0.0) | 1 (0.7) | 0 (0.0) | 1 (0.4) |  |
| **Number of sexual partners in the last 6 months** |  |  |  |  |  |
| None | 0 (0.0) | 1 (0.7) | 1 (1.0) | 2 (0.8) | 0.982 |
| 1 partner | 8 (88.9) | 113 (83.1) | 88 (83.8) | 209 (83.6) |  |
| 2-3 partners | 1 (11.1) | 21 (15.4) | 16 (15.2) | 38 (15.2) |  |
| 4-5 partners | 0 (0.0) | 1 (0.7) | 0 (0.0) | 1 (0.4) |  |
| **Partner's HIV status** |  |  |  |  |  |
| Negative | 4 (44.4) | 79 (58.1) | 49 (46.7) | 132 (52.8) | 0.426 |

|  |  |  |  |  |
| --- | --- | --- | --- | --- |
| Positive | 0 (0.0) | 2 (1.5) | 1 (1.0) | 3 (1.2) |
| Unknown | 5 (55.6) | 55 (40.4) | 55 (52.4) | 115 (46.0) |

**Note:** VOICE risk categories were defined as low risk low risk (0-3 points), moderate risk (4-8 points), and high risk ( $\geq 9$ points)

### Participant characteristics by PRHS categories

Participants were most commonly classified in the moderate PRHS category (62.4%), followed by low (31.6%) and high (6.0%) categories (Table 3). PRHS categories were significantly associated with anal intercourse, oral intercourse, consistent condom use during the past month, ever having sold sex, and age-disparate sexual partnerships (all p<0.05). The proportion reporting these behaviours varied across categories; for example, anal intercourse was reported by 42.9% of participants in the moderate-risk category, compared with 20.3% in the low-risk and 6.7% in the high-risk category. No significant differences were observed in age, education/employment status, household food insecurity, financial access to healthcare, partner characteristics, or partner HIV status.

**Table 3.**
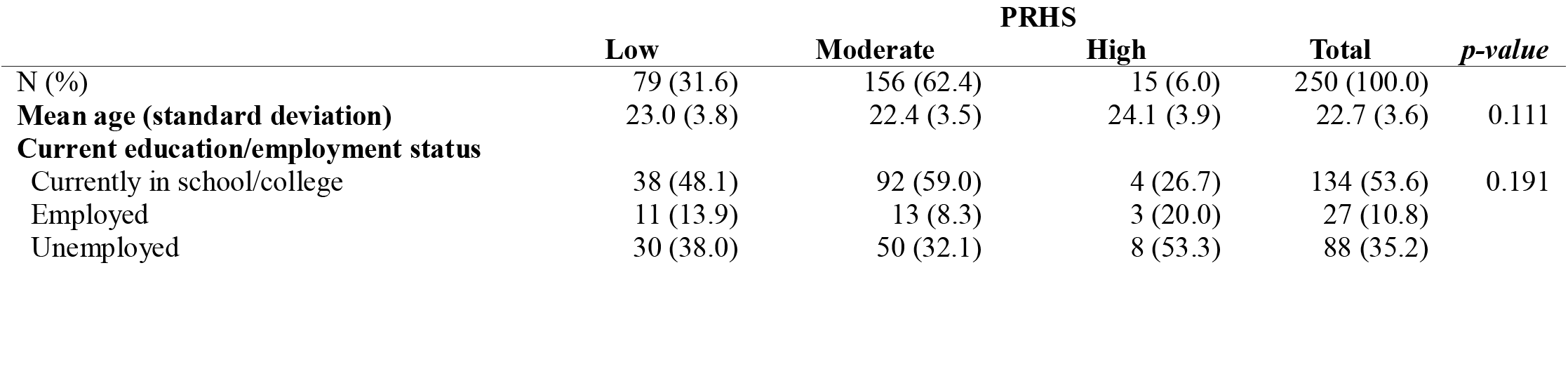
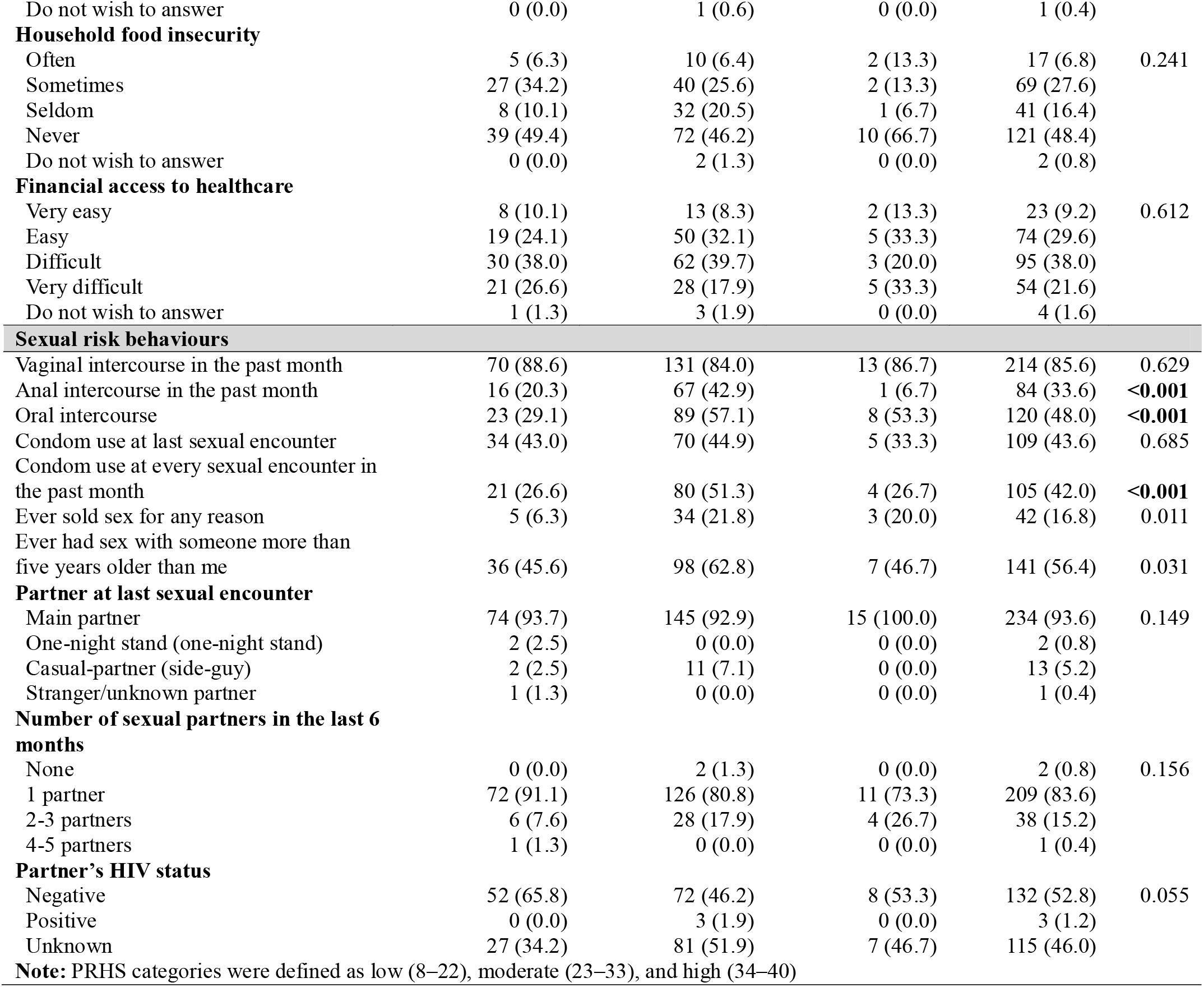
Sociodemographic, household, and sexual risk behaviour characteristics by PRHS.

|  | PRHS |  |  |  |  |
| --- | --- | --- | --- | --- | --- |
|  | Low | Moderate | High | Total | *p-value* |
| N (%) | 79 (31.6) | 156 (62.4) | 15 (6.0) | 250 (100.0) |  |
| Mean age (standard deviation) | 23.0 (3.8) | 22.4 (3.5) | 24.1 (3.9) | 22.7 (3.6) | 0.111 |
| Current education/employment status |  |  |  |  |  |
| Currently in school/college | 38 (48.1) | 92 (59.0) | 4 (26.7) | 134 (53.6) | 0.191 |
| Employed | 11 (13.9) | 13 (8.3) | 3 (20.0) | 27 (10.8) |  |
| Unemployed | 30 (38.0) | 50 (32.1) | 8 (53.3) | 88 (35.2) |  |
| Do not wish to answer | 0 (0.0) | 1 (0.6) | 0 (0.0) | 1 (0.4) |  |
| **Household food insecurity** |  |  |  |  |  |
| Often | 5 (6.3) | 10 (6.4) | 2 (13.3) | 17 (6.8) | 0.241 |
| Sometimes | 27 (34.2) | 40 (25.6) | 2 (13.3) | 69 (27.6) |  |
| Seldom | 8 (10.1) | 32 (20.5) | 1 (6.7) | 41 (16.4) |  |
| Never | 39 (49.4) | 72 (46.2) | 10 (66.7) | 121 (48.4) |  |
| Do not wish to answer | 0 (0.0) | 2 (1.3) | 0 (0.0) | 2 (0.8) |  |
| **Financial access to healthcare** |  |  |  |  |  |
| Very easy | 8 (10.1) | 13 (8.3) | 2 (13.3) | 23 (9.2) | 0.612 |
| Easy | 19 (24.1) | 50 (32.1) | 5 (33.3) | 74 (29.6) |  |
| Difficult | 30 (38.0) | 62 (39.7) | 3 (20.0) | 95 (38.0) |  |
| Very difficult | 21 (26.6) | 28 (17.9) | 5 (33.3) | 54 (21.6) |  |
| Do not wish to answer | 1 (1.3) | 3 (1.9) | 0 (0.0) | 4 (1.6) |  |
| **Sexual risk behaviours** |  |  |  |  |  |
| Vaginal intercourse in the past month | 70 (88.6) | 131 (84.0) | 13 (86.7) | 214 (85.6) | 0.629 |
| Anal intercourse in the past month | 16 (20.3) | 67 (42.9) | 1 (6.7) | 84 (33.6) | <0.001 |
| Oral intercourse | 23 (29.1) | 89 (57.1) | 8 (53.3) | 120 (48.0) | <0.001 |
| Condom use at last sexual encounter | 34 (43.0) | 70 (44.9) | 5 (33.3) | 109 (43.6) | 0.685 |
| Condom use at every sexual encounter in the past month | 21 (26.6) | 80 (51.3) | 4 (26.7) | 105 (42.0) | <0.001 |
| Ever sold sex for any reason | 5 (6.3) | 34 (21.8) | 3 (20.0) | 42 (16.8) | 0.011 |
| Ever had sex with someone more than five years older than me | 36 (45.6) | 98 (62.8) | 7 (46.7) | 141 (56.4) | 0.031 |
| **Partner at last sexual encounter** |  |  |  |  |  |
| Main partner | 74 (93.7) | 145 (92.9) | 15 (100.0) | 234 (93.6) | 0.149 |
| One-night stand (one-night stand) | 2 (2.5) | 0 (0.0) | 0 (0.0) | 2 (0.8) |  |
| Casual-partner (side-guy) | 2 (2.5) | 11 (7.1) | 0 (0.0) | 13 (5.2) |  |
| Stranger/unknown partner | 1 (1.3) | 0 (0.0) | 0 (0.0) | 1 (0.4) |  |
| **Number of sexual partners in the last 6 months** |  |  |  |  |  |
| None | 0 (0.0) | 2 (1.3) | 0 (0.0) | 2 (0.8) | 0.156 |
| 1 partner | 72 (91.1) | 126 (80.8) | 11 (73.3) | 209 (83.6) |  |
| 2-3 partners | 6 (7.6) | 28 (17.9) | 4 (26.7) | 38 (15.2) |  |
| 4-5 partners | 1 (1.3) | 0 (0.0) | 0 (0.0) | 1 (0.4) |  |
| **Partner's HIV status** |  |  |  |  |  |
| Negative | 52 (65.8) | 72 (46.2) | 8 (53.3) | 132 (52.8) | 0.055 |
| Positive | 0 (0.0) | 3 (1.9) | 0 (0.0) | 3 (1.2) |  |
| Unknown | 27 (34.2) | 81 (51.9) | 7 (46.7) | 115 (46.0) |  |

**Note:** PRHS categories were defined as low (8–22), moderate (23–33), and high (34–40)

### Participant characteristics by risk-taking propensity categories

Participants were distributed approximately equally across the three BART risk-taking propensity groups, with 33.5% in both the lowest and middle thirds and 33.1% in the highest third (Table 4). The only statistically significant association with BART category was vaginal intercourse in the past month (p=0.042), reported by 77.8% of participants in the lowest third and 91.2% in the highest third. Age, education/employment status, household food insecurity, financial access to healthcare, other sexual behaviours, partner characteristics, and partner HIV status did not differ significantly across BART categories.

**Table 4.** Sociodemographic, household, and sexual risk behaviour characteristics by Risk-taking propensity.

|  | RISK-TAKING PROPENSITY |  |  |  | p-value |
| --- | --- | --- | --- | --- | --- |
|  | Lowest third | Middle third | Highest third | Total |  |
| N (%) | 81 (33.5) | 81 (33.5) | 80 (33.1) | 242 (100.0) |  |
| Mean age (standard deviation) | 22.7 (3.5) | 22.9 (3.9) | 22.4 (3.5) | 22.7 (3.6) | 0.698 |
| Current education/employment status |  |  |  |  |  |
| Currently in college | 42 (51.9) | 48 (59.3) | 40 (50.0) | 130 (53.7) | 0.720 |
| Employed | 10 (12.3) | 7 (8.6) | 10 (12.5) | 27 (11.2) |  |
| Unemployed | 29 (35.8) | 26 (32.1) | 29 (36.2) | 84 (34.7) |  |
| Do not wish to answer | 0 (0.0) | 0 (0.0) | 1 (1.2) | 1 (0.4) |  |
| **Household food insecurity** |  |  |  |  |  |
| Often | 3 (3.7) | 10 (12.3) | 4 (5.0) | 17 (7.0) | 0.449 |
| Sometimes | 19 (23.5) | 23 (28.4) | 23 (28.7) | 65 (26.9) |  |
| Seldom | 14 (17.3) | 12 (14.8) | 15 (18.8) | 41 (16.9) |  |
| Never | 44 (54.3) | 36 (44.4) | 37 (46.2) | 117 (48.3) |  |
| Do not wish to answer | 1 (1.2) | 0 (0.0) | 1 (1.2) | 2 (0.8) |  |
| **Financial access to healthcare** |  |  |  |  |  |
| Very easy | 9 (11.1) | 8 (9.9) | 5 (6.2) | 22 (9.1) | 0.095 |
| Easy | 30 (37.0) | 21 (25.9) | 21 (26.2) | 72 (29.8) |  |
| Difficult | 25 (30.9) | 26 (32.1) | 40 (50.0) | 91 (37.6) |  |
| Very difficult | 15 (18.5) | 24 (29.6) | 14 (17.5) | 53 (21.9) |  |
| Do not wish to answer | 2 (2.5) | 2 (2.5) | 0 (0.0) | 4 (1.7) |  |
| **Sexual risk behaviours** |  |  |  |  |  |
| Had vaginal sex in the past month | 63 (77.8) | 71 (87.7) | 73 (91.2) | 207 (85.5) | 0.042 |
| Had anal sex in the past month | 30 (37.0) | 23 (28.4) | 27 (33.8) | 80 (33.1) | 0.498 |
| Had oral sex in the past month | 41 (50.6) | 32 (39.5) | 41 (51.2) | 114 (47.1) | 0.243 |
| Condom use at the last sexual encounter | 35 (43.2) | 31 (38.3) | 40 (50.0) | 106 (43.8) | 0.322 |
| Condom use at every sexual encounter in the past month | 35 (43.2) | 28 (34.6) | 37 (46.2) | 100 (41.3) | 0.295 |
| Ever sold sex for any reason | 11 (13.6) | 15 (18.5) | 16 (20.0) | 42 (17.4) | 0.530 |
| Ever had sex with someone who was five years older than me | 50 (61.7) | 48 (59.3) | 38 (47.5) | 136 (56.2) | 0.152 |
| **Partner at last sexual encounter** |  |  |  |  |  |
| Main partner | 74 (91.4) | 76 (93.8) | 76 (95.0) | 226 (93.4) | 0.786 |
| One-time sexual partner (one-night stand) | 1 (1.2) | 1 (1.2) | 0 (0.0) | 2 (0.8) |  |
| Casual-partner (side-guy) | 5 (6.2) | 4 (4.9) | 4 (5.0) | 13 (5.4) |  |
| Stranger/unknown partner | 1 (1.2) | 0 (0.0) | 0 (0.0) | 1 (0.4) |  |
| **Number of sexual partners in the last 6 months** |  |  |  |  |  |
| None | 1 (1.2) | 1 (1.2) | 0 (0.0) | 2 (0.8) | 0.757 |
| 1 partner | 66 (81.5) | 67 (82.7) | 68 (85.0) | 201 (83.1) |  |
| 2-3 partners | 14 (17.3) | 13 (16.0) | 11 (13.8) | 38 (15.7) |  |
| 4-5 partners | 0 (0.0) | 0 (0.0) | 1 (1.2) | 1 (0.4) |  |
| **Partner's HIV status** |  |  |  |  |  |
| Negative | 41 (50.6) | 40 (49.4) | 47 (58.8) | 128 (52.9) | 0.450 |
| Positive | 0 (0.0) | 2 (2.5) | 1 (1.2) | 3 (1.2) |  |
| Unknown | 40 (49.4) | 39 (48.1) | 32 (40.0) | 111 (45.9) |  |

**Note:** Totals differ because analyses involving BART were restricted to participants with complete BART data (n = 242); Risk-taking propensity categories were based on tertiles of the distribution of adjusted average BART pumps: lowest third, middle third, and highest third. These categories were sample-derived and do not represent validated clinical cut-offs. Analyses were restricted to participants with complete BART data (n=242).

### Comparison of participant profiles across the three measures

The three measures showed different patterns of association with participant characteristics and sexual behaviours. VOICE risk categories differed by age and household food insecurity, whereas PRHS categories were associated with five sexual behaviour characteristics namely, anal intercourse, oral intercourse, consistent condom use, ever having sold sex, and age-disparate sexual partnerships. In contrast, BART risk-taking propensity was associated only with vaginal intercourse in the past month. The proportion classified in the high-risk category also differed across measures, at 42.0% for VOICE and 6.0% for PRHS, while 33.1% of participants with complete BART data were in the highest tertile of risk-taking propensity. These descriptive findings suggest that the measures capture different aspects of participant risk profiles rather than identifying identical groups.

### Dissociation of risk measures

Spearman’s rank correlation analyses showed weak, statistically non-significant associations between the three measures (Figure 1). The correlation between BART-adjusted average pumps and PRHS scores was negative and negligible (Spearman’s ρ = 0.050, p = 0.442). The correlation between BART-adjusted average pumps and VOICE risk scores was also negligible (Spearman’s ρ = −0.005, p = 0.941). Similarly, the association between PRHS and VOICE risk scores was weak and positive (Spearman’s ρ = 0.038, p = 0.548).

### Comparative distribution of standardized risk metrics

The frequency distributions of the three standardised HIV risk metrics demonstrate a lack of alignment between behaviour-based HIV risk, self-perceived risk, and general risk-taking propensity (Figure 2). The VOICE score distribution (red) was concentrated towards the higher end of the scale, with the highest frequency of scores between 70% and 80%. However, 42.0% of participants were classified as having high behaviour-based HIV risk (Table 2). The PRHS distribution (green) and BART distribution (blue) showed different patterns, highlighting the distinct dimensions of risk captured by each measure. In contrast, the frequency distribution for self-perceived risk (PRHS; green) is more centrally located, with the largest clusters of participants scoring in the 40% to 75% range. Finally, the frequency of BART scores (blue) is widely distributed across the lower-to-middle range (20-60%), exhibiting the highest variance in the sample.

## Discussion

In this study of young women aged 18-30 in rural KZN, we found limited convergence between risk-taking propensity, self-perceived HIV risk, and self-reported behaviour-based HIV vulnerability. Correlations between risk-taking propensity and both the PRHS and VOICE risk scores were weak and statistically non-significant (p>0.05), as were correlations between the PRHS and VOICE risk score. Our principal finding in this study is that risk-taking propensity is not strongly correlated with either of the two measures. Instead, it suggests that the BART is a separate construct where an individual’s internal drive for reward-seeking and their personal appraisal of HIV vulnerability (PRHS) operate independently from the high-risk sexual behaviours they report (VOICE). This interpretation is further supported by the finding that participants classified into different BART tertiles were remarkably similar across demographic, socioeconomic, and most sexual behavioural characteristics, with recent vaginal intercourse being the only behaviour that differed significantly. The absence of consistent behavioural differences reinforces the possibility that the BART measures a general propensity for risk-taking that is distinct from HIV-specific sexual risk. Although household food insecurity and other structural vulnerabilities were prevalent in this population, food insecurity differed significantly across VOICE risk categories (p=0.029). In contrast, age-disparate sex (p=0.478) and transactional sex (p=0.496) were not significantly associated with VOICE risk category. These findings suggest that the relationship between structural circumstances and reported HIV vulnerability may be complex, and that the factors distinguishing VOICE risk categories in this sample should be interpreted in light of the observed associations rather than assumed relationships between socioeconomic conditions, sexual relationships, and risk.

Consistent with our findings, prior studies in SSA have documented a disconnect between self-reported behavioural HIV risk and self-perceived risk in AGYW. A cross-sectional analysis across DREAMS programme sites in Kenya, Malawi, and Zambia found that almost all sexually active AGYW reported at least one sexual risk factor (>98%) while a much smaller proportion perceived themselves as likely to have been exposed to HIV (Kenya 15.7%; Malawi 46.2%; Zambia 9.5%), highlighting a disconnect between behaviour-based HIV risk and self-perceived risk (43). Similarly, a cross-sectional study of 1,000 sexually active AGYW aged 15-24 years in Malawi found that although having multiple HIV risk factors was associated with increased HIV prevalence, a substantial proportion of those with the highest risk burden did not perceive themselves to be at high HIV risk, 63% of those with ≥5 risk factors and 52% of those with >8 risk factors reported low perceived HIV risk (22).

Despite these weak overall correlations, our findings suggest that self-perceived HIV risk was associated with certain sexual behaviours. High perceived HIV risk was observed alongside some reported sexual behaviours, although patterns were not consistent across all indicators. Associations with the number of sexual partners (p=0.156) and uncertainty regarding partners’ HIV status (p=0.055) did not reach statistical significance. Furthermore, the distribution of some behaviours, including anal intercourse, did not follow a consistent gradient across PRHS categories; the small number of participants reporting anal intercourse in the high-PRHS group may have contributed to this pattern. These findings suggest that self-perceived HIV risk did not consistently correspond with the distribution of individual reported sexual behaviours as measured by the VOICE Risk Score in this sample. Self-perceived HIV risk may therefore capture an important, but incomplete, dimension of HIV vulnerability that complements rather than substitutes for behaviour-based risk assessment.

The limited convergence between behavioural risk-taking propensity, self-perceived HIV risk, and behaviour-based HIV vulnerability observed in this study is plausible given the cognitive, emotional, and sociocultural factors shaping risk appraisal. Research across health domains shows that perceived risk may diverge from behavioural or biomarker-based indicators (44, 45). Behavioural economics suggests that present bias, information salience, and optimism bias may influence how individuals weigh immediate rewards against future health risks, potentially contributing to discrepancies between perceived risk and reported behaviour (46, 47).

Psychosocial factors, including gender norms, relationship dynamics, trust in partners, and anticipated stigma, may further shape young women’s perceptions of HIV risk and engagement with prevention. Skovdal et al. (2022) highlight how expectations surrounding femininity and being a “good girl” or compliant partner can influence young women’s recognition of risk and uptake of prevention options such as pre-exposure prophylaxis (PrEP) (48, 49). South African studies similarly suggest that AGYW may underestimate their HIV risk because of perceived partner fidelity, trust, or the absence of symptoms (20). Together, these findings suggest that self-perceived risk, reported behaviour, and behavioural risk-taking propensity capture related but distinct dimensions of HIV vulnerability and should be interpreted as complementary rather than interchangeable measures.

This study has several strengths, including the use of multiple, conceptually distinct measures within a high-HIV-burden setting in SSA and recruitment within a Health and Demographic Surveillance System (HDSS). Applying the measures within the same sample enabled us to examine the extent to which they identified overlapping or different dimensions of risk. Several limitations should also be considered. First, the bivariate correlation analyses did not account for potential confounding factors, and the cross-sectional design precludes conclusions about causal or temporal relationships. The weak correlations may also reflect dynamic relationships between risk perception, risk-taking propensity, and sexual behaviour that cannot be captured at a single time point. Second, recruitment through the HDSS and *Thetha Nami* safe spaces provided a well-characterized study context but may limit generalizability to young women outside the surveillance area, in urban settings, or who do not access community-based services. Women attending safe spaces may differ in health-seeking behaviour, prevention awareness, or engagement with support services from those who do not attend. The findings should therefore be interpreted in relation to this study population rather than assumed to represent all young women in South Africa.

Third, the BART was administered without tangible financial rewards or penalties. If incentives influence engagement with the task, their absence may have affected its sensitivity to risk-taking propensity. Furthermore, a general behavioural task may not fully capture the relationship-specific and sociocultural contexts in which sexual decisions are made. Fourth, sexual behaviours were self-reported and may be subject to recall and social desirability biases. Finally, the PRHS has not previously been validated in South Africa, which may affect the interpretation and transferability of its scores in this context.

Accurately identifying HIV vulnerability among young women in uMkhanyakude is important in a setting where HIV prevention remains a public health priority. If prevention decisions rely solely on self-perceived risk, young women who report potential HIV exposure but do not consider themselves at risk may be overlooked. Self-reported sexual histories can also be affected by recall, social desirability, and underreporting, reinforcing the need to interpret individual measures cautiously. Prevention programmes should therefore avoid relying exclusively on an individual’s risk appraisal when offering prevention services. Proactive, non-stigmatizing approaches, including routine PrEP counselling and differentiated service delivery, may help ensure that young women are informed about available prevention options. Behaviour-based tools such as the VOICE Risk Score may provide complementary information, but further research is needed to determine whether combining these assessments improves prevention access, uptake, or outcomes. Risk communication should frame HIV vulnerability as situational and cumulative rather than identity-based, avoiding stigmatizing labels that may discourage prevention engagement.

In summary, our findings highlight that risk-taking is a multidimensional construct, with behavioural performance, clinical risk scores, and self-reported perceptions capturing distinct facets of vulnerability. The weak correlations between these domains suggest that being highly vulnerable in one measure does not reliably indicate high vulnerability in another. These findings highlight the complexity of identifying individual-level HIV vulnerability and suggest that integrated, multidimensional risk assessment approaches may be needed, particularly when attempting to identify need for HIV prevention interventions. Future research should explore longitudinal pathways between these constructs and HIV incidence and evaluate whether domain-specific behavioural tasks or culturally tailored instruments better predict HIV vulnerability and, therefore, uptake of prevention behaviours such as PrEP and consistent condom use in high-burden settings.

## Data Availability

All data produced in the present study are available upon reasonable request to the authors

## Author Contributions

Conceptualization – SD, CH, LCL, JM Data Curation – SD, RG, EM, LCL

Formal Analysis – SD, CH, RG, DG, EM, LCL Funding Acquisition – JM, CH, DG, LCL Investigation – SD, CH, DG, JM, EM Methodology – SD, CH, JM, DG

Project Administration – SD, CH, DG, JM Resources – JM, CH, DG, MS

Software – RG, EM Supervision – CH, JM, EM, DG

Validation – SD, CH, JM, LCL, EM Visualization – SD, CH, EM, JM, LCL

Writing – Original draft preparation – SD, CH, JM, DG

Writing – Review and editing – SD, CH, RG, MS, DG, EM, JM, LCL

## Competing Interests

No competing interests were disclosed.

## Grant Information & Disclaimer

This research was nested in a larger study funded by the Gates Foundation (grant number: INV-039268), in which SD assumes the position of a Research Fellow. Additionally, the work reported herein was also made possible through funding by the South African Medical Research Council (SAMRC) through its Division of Research Capacity Development under the Postgraduate Research Associate Programme awarded to SD. Additional funding support was received from the Health Economics and Epidemiology Research (HE^2^RO), SAMRC Health Systems Research Unit and the Youth Health Economics focal area of the SAMRC Health Systems Research Unit. The content hereof is the sole responsibility of the authors and does not necessarily represent the official views of the Gates Foundation, HE^2^RO or SAMRC. DG is funded by the SAMRC/Wits Centre for Health Economics and Decision Science – PRICELESS SA (Grant number 23108).

## Conflict of Interest

The authors have no conflicts of interest.

## Notes

### Competing Interest Statement

The authors have declared no competing interest.

### Author Declarations

University of the Witwatersrand Human Research Ethics Committee KwaZulu Natal Provincial Department of Health uMkhanyakude Health District

## References

1. UNAIDS. Global HIV & AIDS staistics - Fact Sheet 2025. Geneva, Switzerland; 2025.

2. Chimbindi N, Birdthistle I, Floyd S, Harling G, Mthiyane N, Zuma T, et al. Directed and target focused multi-sectoral adolescent HIV prevention: Insights from implementation of the ‘DREAMS Partnership’ in rural South Africa. J Int AIDS Soc. 2020;23 Suppl 5(Suppl 5):e25575.

3. Chimbindi N, Birdthistle I, Shahmanesh M, Osindo J, Mushati P, Ondeng’e K, et al. Translating DREAMS into practice: Early lessons from implementation in six settings. PLOS ONE. 2018;13(12):e0208243.

4. UNAIDS. The Global AIDS Update 2024 2024.

5. Lavoie M-CC, Blanco N, Lascko T, Baumhart C, Verbeek J, Stoebenau K, et al. The effects of the Determined, Resilient, Empowered, AIDS-free, Mentored, and Safe (DREAMS) program on HIV incidence and other health-related outcomes among adolescent girls and young women: a systematic review and meta-analysis. eClinicalMedicine. 2025;82.

6. Reitsema M, Wallinga J, van Sighem AI, Bezemer D, van der Valk M, van Aar F, et al. Impact of Varying Pre-exposure Prophylaxis Programs on HIV and Neisseria gonorrhoeae Transmission Among MSM in the Netherlands: A Modeling Study. JAIDS Journal of Acquired Immune Deficiency Syndromes. 2024;97(4):325–33.

7. Hendrickson C, Hirasen K, Mongwenyana C, Benade M, Bothma R, Smith C, et al. Costs and outcomes of routine HIV oral pre-exposure prophylaxis implementation across different service delivery models and key populations in South Africa: a retrospective cohort study. The Lancet HIV. 2025;12(2):e130–e42.

8. Lewis L, Kharsany ABM, Humphries H, Maughan-Brown B, Beckett S, Govender K, et al. HIV incidence and associated risk factors in adolescent girls and young women in South Africa: A population-based cohort study. PLoS One. 2022;17(12):e0279289.

9. Cassim N, Coetzee LM, da Silva MP, Glencross DK, Stevens WS. Assessing very advanced HIV disease in adolescent girls and young women. South Afr J HIV Med. 2023;24(1):1501.

10. Jewkes RK, Dunkle K, Nduna M, Shai N. Intimate partner violence, relationship power inequity, and incidence of HIV infection in young women in South Africa: a cohort study. The Lancet. 2010;376(9734):41–8.

11. Siemieniuk RA, Krentz HB, Gish JA, Gill MJ. Domestic violence screening: prevalence and outcomes in a Canadian HIV population. AIDS Patient Care STDS. 2010;24(12):763–70.

12. Sullivan TP. The intersection of intimate partner violence and HIV: detection, disclosure, discussion, and implications for treatment adherence. Top Antivir Med. 2019;27(2):84–7.

13. Stoner MCD, Pettifor A, Edwards JK, Aiello AE, Halpern CT, Julien A, et al. The effect of school attendance and school dropout on incident HIV and HSV-2 among young women in rural South Africa enrolled in HPTN 068. Aids. 2017;31(15):2127–34.

14. Maje L. Thesis on the association of vaginal practices to bacterial vaginosis among adolescent girls and young women in South Africa: A risk for HIV. 2019.

15. Kassanjee R, McWalter TA, Bärnighausen T, Welte A. A new general biomarker-based incidence estimator. Epidemiology. 2012;23(5):721–8.

16. Pettifor AE, Van der Straten A, Dunbar MS, Shiboski SC, Padian NS. Early age of first sex: a risk factor for HIV infection among women in Zimbabwe. Aids. 2004;18(10):1435–42.

17. Lewis JJ, Donnelly CA, Mare P, Mupambireyi Z, Garnett GP, Gregson S. Evaluating the proximate determinants framework for HIV infection in rural Zimbabwe. Sexually transmitted infections. 2007;83(suppl 1):i61–i9.

18. Sambisa W, Curtis SL, Stokes CS. Ethnic differences in sexual behaviour among unmarried adolescents and young adults in Zimbabwe. Journal of biosocial science. 2010;42(1):1–25.

19. Machemedze T. Does self-perceived HIV risk mediate the potential association between HIV-related symbolic stigma and sexual behaviour among young adult women in Cape Town, South Africa? BMC Public Health. 2023;23(1):188.

20. Maughan-Brown B, Venkataramani AS. Accuracy and determinants of perceived HIV risk among young women in South Africa. BMC Public Health. 2017;18(1):42.

21. Kamire V, Magut F, Khagayi S, Kambona C, Muttai H, Nganga L, et al. HIV Risk Factors and Risk Perception Among Adolescent Girls and Young Women: Results From a Population-Based Survey in Western Kenya, 2018. J Acquir Immune Defic Syndr. 2022;91(1):17–25.

22. Price JT, Rosenberg NE, Vansia D, Phanga T, Bhushan NL, Maseko B, et al. Predictors of HIV, HIV Risk Perception, and HIV Worry Among Adolescent Girls and Young Women in Lilongwe, Malawi. J Acquir Immune Defic Syndr. 2018;77(1):53–63.

23. Jia KM, Eilerts H, Edun O, Lam K, Howes A, Thomas ML, et al. Risk scores for predicting HIV incidence among adult heterosexual populations in sub-Saharan Africa: a systematic review and meta-analysis. Journal of the International AIDS Society. 2022;25(1):e25861.

24. Oo MM, Rudd M, Shukalek C, Kishibe T, Hull M, Tan DHS. Diagnostic performance of HIV risk assessment tools for identifying pre-exposure prophylaxis candidates: a systematic review and meta-analysis. eClinicalMedicine. 2025;88:103487.

25. Carroll JJ, Heffron R, Mugo N, Ngure K, Ndase P, Asiimwe S, et al. Perceived Risk Among Human Immunodeficiency Virus Serodiscordant Couples in East Africa Taking Oral Pre-Exposure Prophylaxis. Sexually Transmitted Diseases. 2016;43(8).

26. Derefinko KJ, Peters JR, Eisenlohr-Moul TA, Walsh EC, Adams ZW, Lynam DR. Relations between trait impulsivity, behavioral impulsivity, physiological arousal, and risky sexual behavior among young men. Arch Sex Behav. 2014;43(6):1149–58.

27. Walters KJ, Webb MK, Simons JS. The Implicit Sexual Risk Assessment: A Pilot Study of a Novel Behavioral Task. Arch Sex Behav. 2022;51(6):2921–9.

28. Rose AK, Jones A, Clarke N, Christiansen P. Alcohol-induced risk taking on the BART mediates alcohol priming. Psychopharmacology (Berl). 2014;231(11):2273–80.

29. Gareta D, Baisley K, Mngomezulu T, Smit T, Khoza T, Nxumalo S, et al. Cohort Profile Update: Africa Centre Demographic Information System (ACDIS) and population-based HIV survey. Int J Epidemiol. 2021;50(1):33–4.

30. Kharsany ABM, Cawood C, Khanyile D, Lewis L, Grobler A, Puren A, et al. Community-based HIV prevalence in KwaZulu-Natal, South Africa: results of a cross-sectional household survey. Lancet HIV. 2018;5(8):e427–e37.

31. Busang J, Zuma T, Herbst C, Okesola N, Chimbindi N, Dreyer J, et al. Thetha Nami ngithethe nawe (Let’s Talk): a stepped-wedge cluster randomised trial of social mobilisation by peer navigators into community-based sexual health and HIV care, including pre-exposure prophylaxis (PrEP), to reduce sexually transmissible HIV amongst young people in rural KwaZulu-Natal, South Africa. BMC Public Health. 2023;23(1):1553.

32. Shahmanesh M, Okesola N, Chimbindi N, Zuma T, Mdluli S, Mthiyane N, et al. Thetha Nami: participatory development of a peer-navigator intervention to deliver biosocial HIV prevention for adolescents and youth in rural South Africa. BMC Public Health. 2021;21(1):1393.

33. Harris PA, Taylor R, Thielke R, Payne J, Gonzalez N, Conde JG. Research electronic data capture (REDCap)—a metadata-driven methodology and workflow process for providing translational research informatics support. Journal of biomedical informatics. 2009;42(2):377–81.

34. Busang J, Ngoma N, Zuma T, Herbst C, Okesola N, Chimbindi N, et al. Person-centred HIV care and prevention for youth in rural South Africa: preliminary implementation findings from Thetha Nami ngithethe nawe stepped-wedge trial of peer-navigator mobilization into mobile sexual health services. Journal of the International AIDS Society. 2025;28(S5):e70032.

35. Chen DL, Schonger M, Wickens C. oTree—An open-source platform for laboratory, online, and field experiments. Journal of Behavioral and Experimental Finance. 2016;9:88–97.

36. Balkus JE, Brown E, Palanee T, Nair G, Gafoor Z, Zhang J, et al. An Empiric HIV Risk Scoring Tool to Predict HIV-1 Acquisition in African Women. J Acquir Immune Defic Syndr. 2016;72(3):333–43.

37. Rosenberg NE, Kudowa E, Price JT, Pettifor A, Bekker LG, Hosseinipour MC, et al. Identifying Adolescent Girls and Young Women at High Risk for HIV Acquisition: A Risk Assessment Tool From the Girl Power-Malawi Study. Sex Transm Dis. 2020;47(11):760–6.

38. Napper LE, Fisher DG, Reynolds GL. Development of the perceived risk of HIV scale. AIDS Behav. 2012;16(4):1075–83.

39. Lukhele BW, Musumari P, El-Saaidi C, Techasrivichien T, Suguimoto SP, Ono Kihara M, et al. Efficacy of Mobile Serious Games in Increasing HIV Risk Perception in Swaziland: A Randomized Control Trial (SGprev Trial) Research Protocol. JMIR Res Protoc. 2016;5(4):e224.

40. Onyechi KCN, Eseadi C, Okere AU, Otu MS. Effects of Rational-Emotive Health Education Program on HIV risk perceptions among in-school adolescents in Nigeria. Medicine (Baltimore). 2016;95(29):e3967.

41. Lejuez CW, Read JP, Kahler CW, Richards JB, Ramsey SE, Stuart GL, et al. Evaluation of a behavioral measure of risk taking: The Balloon Analogue Risk Task (BART). Journal of Experimental Psychology: Applied. 2002;8(2):75–84.

42. StataCorp. Stata Statistical Software: Release 19. 19.5 ed. College Station, TX2025.

43. Heck CJ, Reed DM, Okal J, Chipeta E, Mbizvo M, Mathur S. Examining concordance of sexual-related factors and PrEP eligibility with HIV risk perception among adolescent girls and young women: cross-sectional insights from DREAMS sites in Kenya, Malawi, and Zambia. BMC Public Health. 2024;24(1):2793.

44. Brawarsky P, Eibensteiner K, Klinger EV, Baer HJ, Getty G, Orav EJ, et al. Accuracy of self-perceived risk for common conditions. Cogent Med. 2018;5.

45. Rodriguez SA, Tiro JA, Baldwin AS, Hamilton-Bevil H, Bowen M. Measurement of Perceived Risk of Developing Diabetes Mellitus: A Systematic Literature Review. J Gen Intern Med. 2023;38(8):1928–54.

46. Linnemayr S, Stecher C. Behavioral Economics Matters for HIV Research: The Impact of Behavioral Biases on Adherence to Antiretrovirals (ARVs). AIDS Behav. 2015;19(11):2069–75.

47. Jennings Mayo-Wilson L, Coleman Lewis J, MacCarthy S, Linnemayr S. Assessing behavioral economic biases among young adults who have increased likelihood of acquiring HIV: a mixed methods study in Baltimore, Maryland. AIDS Research and Therapy. 2023;20(1):25.

48. Skovdal M, Clausen CL, Magoge-Mandizvidza P, Dzamatira F, Maswera R, Nyamwanza RP, et al. How gender norms and ‘good girl’ notions prevent adolescent girls and young women from engaging with PrEP: qualitative insights from Zimbabwe. BMC Women’s Health. 2022;22(1):344.

49. Govender K, Beckett S, Mukuku O, Cawood C, George G, Khanyile D, et al. Psychosocial and behavioral factors associated with HIV among adolescent girls and young women in DREAMS districts in South Africa: cross-sectional survey. Front Public Health. 2025;13:1620084.

